# Media Exposure and the Social Determinants of Health Insurance Coverage in Ethiopia 2011-2016

**DOI:** 10.1101/2021.09.07.21263212

**Authors:** Eniola A. Olatunji, Sanam Maredia, Natalie Freeman, Allen Nguyen, David J Washburn

## Abstract

**Background:** In a push for universal health coverage, Ethiopia introduced two insurance schemes in 2010. Yet coverage rates remain very low. To encourage greater adoption, policymakers require a better understanding of who chooses to enroll and which promotional efforts are most effective in encouraging enrollment.

**Objective:** Using nationally representative Demographic and Health Surveys, this research assessed the social determinants of health insurance coverage, including media exposure, in Ethiopia from 2011-2016.

**Methods:** This research analyzed health insurance coverage and other sociodemographic and media exposure variables using multivariable logistic regression model.

**Results:** Health insurance coverage increased 3.30 times from 1.48% in 2011 to 4.89% in 2016. In both years, coverage was associated with higher education, older age, higher wealth levels, and exposure to newspaper and television. Compared to those with no exposure to newspaper, those with newspaper exposure at least once a week were 1.80 times (2011) and 1.86 times (2016) more likely to be insured. Similar results were obtained for television exposure.

**Conclusion:** Initiatives that target the poor and less educated will be necessary if Ethiopia is to achieve universal health coverage. This research suggests that, to date, newspaper and television mediums have been effective promotion mechanisms for growing enrollment.

## Background

In 2014, the United Nations released a list of 17 Sustainable Development Goals (SDG), including the third goal to “ensure healthy lives and promote well-being for all at all ages” (United Nations, 2019). An important part of this goal focuses on achieving universal health coverage (UHC) (United Nations, 2014). UHC includes “financial risk protection, access to quality essential health-care services and access to safe, effective, quality and affordable essential medicines and vaccines for all” (United Nations, 2014). Ethiopia is one of the many countries that has made a commitment to achieve UHC by 2030.

Being the second largest population in sub-Saharan Africa with approximately 108 million people, it is even more important for Ethiopia to provide health coverage for its residents Central Intelligence Agency [CIA], 2019). Majority of the population resides in rural areas, and the economy is largely dependent on agriculture (World Bank Group, 2016). Approximately 80% of morbidity across the country is the result of nutritional and preventable diseases associated with low socioeconomic status (Ali, 2014). Ethiopia suffers from a large communicable disease burden, including problems with tuberculosis, malaria, and HIV (Chaya, 2007). Yet Ethiopia has found success in accomplishing some of the Millennium Development Goals, the precursors to the SDG. Between 1990 and 2015, there was a 67% reduction in under-five mortality to 64 per 1,000 live births, a 50% decline in mortality due to tuberculosis, and a 73% decrease in death caused by malaria (Assefa et al., 2017; CIA, 2019; United Nations, 2019). Life expectancy has increased from 47 years of age in 1990 to 68 in 2020, and the maternal mortality rate per 100,000 live births has been reduced from 1400 in 1990 to 401 in 2017 (Assefa et al., 2017; CIA, 2019; United Nations, 2019). As certain sectors of Ethiopia develop further, non-infectious diseases such as hypertension, cancer, and heart diseases are gradually becoming more critical, especially for the middle and upper classes (Chaya, 2007). Yet, not unlike many other developing settings, while some individuals are able to benefit from the rapidly growing Ethiopian economy, others remain behind, and health disparities are becoming more pronounced.

In 2017, Ethiopia’s Dr. Tedros Adhanom was elected Director-General of the World Health Organization (WHO), and reported that UHC was his top priority, an ambitious goal especially for lower income countries, some of which had been trying to advance UHC for years already (Lavers, 2019). Prior to 2010, before Ethiopia instituted two new types of public insurance, insurance accounted for only 1.5% of the total private health expenditure (Haile et al., 2014). Private insurance, donors, government funding, and out-of-pocket charges had been the main mechanisms for financing health (Haile et al., 2014). In 2010, a newly implemented mandatory Social Health Insurance (SHI) for the formal sector was meant to insure over 10% of the population, and a voluntary Community-Based Health Insurance (CBHI) was proposed to cover over 80% of the population. The SHI would cover many of the formally employed population and their family members. Examples included public servants, permanent employees working in large private organizations, and eligible pensioners (Gidey et al., 2019). Yet, enrollment rates remain very low as the SHI has not achieved coverage of the over 10% as initially intended. A particular challenge of the SHI is enrolling the poor. According to a 2015 report, of the poor who are part of the formal sector and eligible for SHI, only 2% were enrolled (Global Financing Facility, 2019). Also, despite the SHI being mandatory, institutions do not automatically register their eligible members. Without automatic enrollment, potential SHI enrollees may be without insurance coverage. Voluntary CBHI, on the other hand, scaled up nationally five years later in 161 districts after being piloted in 13 districts in the regions of Amhara, Oromia, Tigray, and SNNP (Southern Nations, Nationalities, and Peoples) (Atafu & Kwon, 2018; Federal Democratic Republic of Ethiopia Ethiopian Health Insurance Agency, 2015; Haile et al., 2014).

In an additional 2015 health reform, the Ethiopian Ministry of Health implemented a Health Sector Transformation Plan with numerous goals “to improve equity, coverage and utilization of essential health services, improve quality of health care, and enhance the implementation capacity of the health sector at all levels of the system” (Federal Democratic Republic, 2015). First, to combat a shortage of health providers especially in rural areas, the government sought to train health extension workers that work directly with individual households and at health posts throughout communities (Assefa et al., 2013). Second, a Pharmaceutical Fund and Supply Agency was established to provide essential medical equipment, supplies, and pharmaceuticals (Workie & Ramana, 2013). Third, CBHI was promoted even further, particularly to help cover the poor and those living in rural areas (Jutting, 2004; Mebratie et al., 2015). According to a 2015/2016 Household Health Service Utilization and Expenditure Survey in Ethiopia, 96% of those that were covered by any health insurance were enrolled in CBHI (Federal Democratic Republic, 2017). Furthermore, 97% of the insured respondents were from the four regions that had implemented CBHI (Federal Democratic Republic, 2017). Thus, it is likely that overall health insurance coverage expanded from about 1% to 7% from the 2010/11 to 2015/16 survey was largely due to the expansion of CBHI (Federal Democratic Republic, 2017). The 2015/2016 rate of 7% is slightly higher than what was found in the DHS survey (4.89%) which has a sample size nearly three times larger. It is uncertain what leads to this difference. Regardless, the Household Health Service Utilization and Expenditure Survey did not analyze the determinants of health insurance in a multivariable manner. This work contributes a number of additional bivariate relationships between insurance coverage as well as a controlled multivariable analysis of the determinants of health insurance, including exposure to various media sources.

Critical to expanding health care coverage in Ethiopia is assessing and understanding who has been left uncovered and why, which is why this work adds critical value. Identifying and analyzing specific social determinants is crucial to provide insight into marginalized groups who need to be specifically targeted for inclusion. The WHO defines the social determinants of health as “the conditions in which people are born, grow, live, work, and age. These circumstances are shaped by the distribution of money, power and resources at global, national and local levels” (Viner et al., 2012). These factors can influence the likelihood that they have access to health insurance, a relationship well-established in the literature (Allcock et al., 2019; Dake, 2018; Kansanga et al., 2018; Kebede et al., 2020; Kimani et al., 2014; Lailulo et al., 2015).

Furthermore, health promotion through the media has been shown to affect health seeking behaviors and decisions to enroll in insurance. For example, research has shown that media exposure through radio, newspaper, and television affects health seeking behaviors including antenatal visits and the initiation of visits in the first trimester in countries in sub-Saharan Africa, including Ethiopia (Okedo-Alex et al., 2019). The relationship between access to different sources of media and enrollment in voluntary insurance schemes has been established in sub-Saharan Africa (Kansanga et al., 2018; Kebede et al., 2020; Kimani et al., 2014). In Kenya, exposure to media was associated with twice the odds of being insured (Kazungu & Barasa, 2017; Kimani et al., 2014). Women who were frequently exposed to radio, read the newspaper, and watched television were highly likely to have health insurance (Kazungu & Barasa, 2017; Kimani et al., 2014). In Ghana, exposure to radio, newspaper, and television was associated with enrollment in the National Health Insurance Scheme. Among Ethiopian women, it was found that reading the newspaper at least once a week was associated with being covered by health insurance (Kebede et al., 2016). The last study only focused on women with the 2016 Demographic and Health Survey. This work expands on that study by identifying those factors that are most associated with health insurance coverage for both men and women in Ethiopia through a multiyear analysis. Critically, this analysis can better inform policymakers about the determinants for coverage for all adults, and how those associations have shifted from the early years of insurance program implementation.

## Methods

The Ethiopia Demographic and Health Surveys (EDHS) for 2011 and 2016 were used for the study (Central Statistical Agency, 2011 & 2016). The EDHS is a nationally representative survey based on multi-stage cluster sampling units. In 2011, the sample included 14,110 males (ages 15-59) and 16,515 females (ages 15-49). In 2016, the sample included 12,688 males (ages 15-59) and 15,683 females (ages 15-49).

Each individual with a response to the question “do you have health insurance coverage?” was included in the analysis. Age was grouped into five categories of ten-year spans with one group for those 45 and older. Occupation was categorized as unemployed, professional, agricultural, and manual labor. Marital status was classified as never married, currently married, living with partner, and formerly/ever married. Wealth was defined by quintiles as poor, poorer, middle, rich, and richest. Exposure to the three media sources newspaper, radio, and television was categorized as not at all (no exposure), exposure less than once a week, and at least once a week.

Descriptive statistics, Pearson chi-square tests for bivariate analyses, and multivariable logistic regressions were used to analyze the data. Bivariate analyses and logistic regression were conducted separately for 2011 and 2016. SAS 9.4 was used for merging the datasets, and STATA 14 was used for the statistical analyses.

## Results

Ultimately, the analysis included 16,501 females and 14,091 males in 2011 and 15,683 females and 12,688 males in 2016. Table 1 shows frequencies and the bivariate analysis for each of the variables. Health insurance coverage was 1.48% in 2011, increasing to 4.89% in 2016. In both years, health insurance coverage was associated with higher education levels, older age, professional and manual employment compared to unemployed, greater wealth, urban residence, and more newspaper and TV exposure. As shown in Table 1, in the bivariate analysis, nearly all the variables except for radio exposure were significant in both 2011 and 2016.

**Table 1.** Bivariate analysis of health insurance coverage in Ethiopia in 2011 and 2016

| Variables | Categories | Total [%]<br>2011 | Number<br>Insured<br>2011 | Percentage<br>insured %<br>2011 | <sup>1</sup>P-values<br>2011 | Total [%]<br>2016 | Number<br>Insured<br>2016 | Percentage<br>insured %<br>2016 | P-values<br>2016 |
| --- | --- | --- | --- | --- | --- | --- | --- | --- | --- |
| Covered by health insurance | No | 30,139<br>[98.52%] |  | 98.52 |  | 26,983<br>[94.95%] |  | 95.10 |  |
|  | Yes | 453<br>[1.48%] |  | 1.84 |  | 1,388<br>[4.89%] |  | 4.90 |  |
| Gender | Female | 16,501<br>[53.94%] | 147 | 0.90 | <0.001 | 15,683<br>[55.28%] | 659 | 4.20 | <0.001 |
|  | Male | 14,091<br>[46.06%] | 306 | 2.20 |  | 12,688<br>[44.72%] | 729 | 5.60 |  |
| Type of residence | Urban | 9,533<br>[31.16%] | 348 | 3.70 | <0.001 | 9,214<br>[32.48%] | 475 | 5.20 | <0.001 |
|  | Rural | 21,059<br>[68.84%] | 105 | 0.50 |  | 19,157<br>[67.52%] | 909 | 4.70 |  |
| Education | No education | 12,712<br>[41.55%] | 31 | 0.20 | <0.001 | 10,550<br>[37.18%] | 472 | 4.50 | <0.001 |
|  | Primary | 12,513<br>[40.90%] | 144 | 1.20 |  | 10,554<br>[37.19%] | 426 | 4.00 |  |
|  | Secondary | 3,019 [9.87%] | 106 | 3.50 |  | 4,471<br>[15.76%] | 222 | 5.00 |  |
|  | Higher | 2,348 [7.68%] | 172 | 7.30 |  | 2,801<br>[9.87%] | 268 | 10.00 |  |
| Age group in years | 15-24 | 12,005<br>[39.24%] | 93 | 0.80 | <0.001 | 10,903<br>[38.43%] | 429 | 3.90 | <0.001 |
|  | 25-34 | 9,231<br>[30.17%] | 183 | 2.00 |  | 8,701<br>[30.67%] | 373 | 4.30 |  |
|  | 35-44 | 6,057<br>[19.80%] | 113 | 1.90 |  | 5,811<br>[20.48%] | 363 | 6.30 |  |
|  | 45 and above | 3,299<br>[10.78%] | 64 | 2.00 |  | 2,956<br>[10.42%] | 223 | 7.50 |  |
| Marital Status | Never married | 10,057<br>[32.87%] | 144 | 1.43 | <0.001 | 9,383<br>[33.07%] | 434 | 4.60 | <0.001 |
|  | Currently married | 16,999<br>[55.57%] | 254 | 1.49 |  | 16,610<br>[58.55%] | 834 | 5.00 |  |
|  | Living with partner | 1,115 [3.64%] | 24 | 2.15 |  | 422 | 19 | 4.50 |  |

<sup>1</sup> \*\*\* = p < 0.001
\*\* = p < 0.01
\* = p < 0.05

|  |  |  |  |  |  |  |  |  |  |
| --- | --- | --- | --- | --- | --- | --- | --- | --- | --- |
|  |  |  |  |  |  | [1.49%] |  |  |  |
|  | Formerly/ever married | 2,421 [7.91%] | 31 | 1.28 |  | 1,956 [6.89%] | 111 | 5.70 |  |
| Occupation | Unemployed | 9,169 [30.25%] | 50 | 0.60 | <0.001 | 9,635 [35.37%] | 212 | 2.20 | <0.001 |
|  | Professional | 17,728 [58.49%] | 240 | 1.40 |  | 6,012 [22.07%] | 360 | 6.00 |  |
|  | Agricultural | 582 [1.92%] | 30 | 5.20 |  | 9,167 [33.65%] | 604 | 6.60 |  |
|  | Manual | 2,833 [9.35%] | 129 | 4.60 |  | 2,429 [8.92%] | 152 | 6.30 |  |
| Wealth | Poorest | 6,442 [21.13%] | 6 | 0.10 | <0.001 | 6,802 [23.97%] | 118 | 1.70 | <0.001 |
|  | Poorer | 4,506 [14.78%] | 3 | 0.10 |  | 3,856 [13.59%] | 182 | 4.70 |  |
|  | Middle | 4,415 [14.48%] | 12 | 0.30 |  | 3,741 [13.19%] | 281 | 7.50 |  |
|  | Rich | 4,904 [16.09] | 29 | 0.60 |  | 3,967 [13.98%] | 276 | 6.70 |  |
|  | Richest | 10,215 [33.51%] | 403 | 4.00 |  | 10,005 [35.26%] | 531 | 5.30 |  |
| Frequency of Newspaper | Not at all | 21,314 [69.78%] | 120 | 0.60 | <0.001 | 21,978 [77.47%] | 913 | 4.20 | <0.001 |
|  | Less than once a week | 6,330 [20.72%] | 175 | 2.80 |  | 4,347 [15.32%] | 272 | 6.30 |  |
|  | At least once a week | 2,901 [9.50%] | 157 | 5.40 |  | 2,046 [7.21%] | 203 | 10.00 |  |
| Frequency of Radio | Not at all | 11,384 [37.25%] | 59 | 0.50 | <0.001 | 16,187 [57.05%] | 662 | 4.10 | <0.001 |
|  | Less than once a week | 9,738 [31.86%] | 136 | 1.40 |  | 5,690 [20.05%] | 271 | 4.80 |  |
|  | At least once a week | 9,439 [30.89%] | 258 | 2.70 |  | 6,496 [22.98%] | 455 | 7.00 |  |
| Frequency of Television | Not at all | 14,140 [46.26%] | 34 | 0.20 | <0.001 | 16,299 [57.45%] | 646 | 4.00 | <0.001 |
|  | Less than once a week | 8,723 [28.54%] | 113 | 1.30 |  | 4,820 [16.99%] | 311 | 6.50 |  |
|  | At least once a week | 7,701 [25.20%] | 304 | 4.00 |  | 7,252 [25.56%] | 431 | 6.00 |  |

Logistic regression models (Table 2) included all the variables from the bivariate analyses. In 2011, when the insurance rate was lower, rural residents were 1.8 times as likely to have health insurance compared to urban residents, and this was statistically significant (OR=1.82; p< 0.001). This difference was no longer significant in 2016 (OR=1.14; p=0.274). Males were statistically significantly more likely to be insured in 2011 (OR=1.42; p<0.004), but this changed in 2016 where males were statistically significantly less likely to be covered by health insurance (OR=0.75; p<0.001).

**Table 2.** Determinants of health insurance adjusted odds ratio with confidence intervals in Ethiopia in 2011 and 2016

| Variable |  | <sup>1</sup>Adjusted Odds Ratio (2011) | Lower, Upper 95% CI (2011) | Adjusted Odds Ratio (2016) | Lower, Upper 95% CI (2016) |
| --- | --- | --- | --- | --- | --- |
| Gender | Female |  |  |  |  |
|  | Male | 1.42** | 1.12, 1.80 | 0.75*** | 0.66, 0.86 |
| Type of Residence | Urban |  |  |  |  |
|  | Rural | 1.82*** | 1.33, 2.49 | 1.14 | 0.90, 1.46 |
| Education | No education |  |  |  |  |
|  | Primary | 2.10*** | 1.37, 3.24 | 0.76*** | 0.65, 0.89 |
|  | Secondary | 3.03*** | 1.87, 4.91 | 1.00 | 0.80, 1.23 |
|  | Higher | 5.61*** | 3.45, 9.10 | 1.93*** | 1.52, 2.44 |
| Age group in years | 15-24 |  |  |  |  |
|  | 25-34 | 1.96*** | 1.47, 2.62 | 0.94 | 0.79, 1.12 |
|  | 35-44 | 2.30*** | 1.63, 3.25 | 1.47*** | 1.21, 1.78 |
|  | 45 and above | 2.63*** | 1.76, 3.94 | 1.68*** | 1.34, 2.09 |
| Marital Status | Never married |  |  |  |  |
|  | Currently married | 1.52** | 1.17, 1.98 | 0.89 | 0.75, 1.05 |
|  | Living with partner | 1.23 | 0.76, 1.97 | 0.70 | 0.42, 1.14 |
|  | Formerly/ever married | 1.02 | 0.66, 1.58 | 1.05 | 0.81, 1.35 |
| Occupation | Unemployed |  |  |  |  |
|  | Professional | 1.78*** | 1.26, 2.49 | 1.93*** | 1.59, 2.33 |
|  | Agricultural | 1.10 | 0.67, 1.78 | 3.19*** | 2.65, 3.84 |
|  | Manual | 3.26*** | 2.25, 4.71 | 2.62*** | 2.09, 3.31 |
| Wealth | Poorest |  |  |  |  |
|  | Poorer | 0.67 | 0.17, 2.68 | 2.43*** | 1.91, 3.10 |
|  | Middle | 2.47 | 0.92, 6.62 | 4.00*** | 3.18, 5.02 |

<sup>1</sup> \*\*\* = p < 0.001
\*\* = p < 0.01
\* = p < 0.05

|  |  |  |  |  |  |
| --- | --- | --- | --- | --- | --- |
|  | Rich | 4.34*** | 1.78, 10.57 | 3.65*** | 2.90, 4.60 |
|  | Richest | 17.10*** | 7.12, 41.03 | 2.37*** | 1.75, 3.19 |
| Frequency of Newspaper | Not at all |  |  |  |  |
|  | Less than once a week | 1.34* | 1.01, 1.76 | 1.27** | 1.07, 1.50 |
|  | At least once a week | 1.80*** | 1.32, 2.44 | 1.86*** | 1.52, 2.27 |
| Frequency of Radio | Not at all |  |  |  |  |
|  | Less than once a week | 1.26 | 0.91, 1.75 | 0.86 | 0.73, 1.01 |
|  | At least once a week | 0.93 | 0.67, 1.28 | 1.08 | 0.92, 1.26 |
| Frequency of Television | Not at all |  |  |  |  |
|  | Less than once a week | 1.71* | 1.12, 2.59 | 1.43*** | 1.21, 1.68 |
|  | At least once a week | 1.83** | 1.19, 2.82 | 1.25* | 1.03, 1.53 |

In 2011, when compared to those with no education, those with higher levels of education had progressively higher odds of health insurance coverage. In comparison to no formal education, higher education had an OR of 5.61 (p<0.001), secondary education had an OR of 3.03 (p<0.001), and primary education had an OR of 2.10 (p<0.001). The result was dissimilar in 2016. Even though higher education had nearly twice the odds of coverage compared to those with no formal education (OR=1.93; p<0.001), secondary education did not have higher odds and was no longer statistically significant (OR=1.00; p<0.973). Primary education was associated with statistically significantly lower odds of health insurance when compared to those with no formal education (OR=0.76; p<0.001).

There was a gradual increase in statistically significant odds ratios with increasing age in 2011 when compared to the reference group (15-24 years). The 25-34 years old group had an OR of 1.96 (p<0.001), and the 35-44 years old group had an OR of 2.30 (p<0.001). The 45 and above age group had an OR of 2.63 (p<0.001). In 2016, only those 35-44 years (OR=1.47; p<0.001) and 45 years and above (OR=1.68; p<0.001) were more likely to be insured than those aged 15-24 years. Only in 2011 was currently being married associated with significantly higher odds of having health insurance compared to people who were never married (OR=1.52; p<0.002).

In 2011, professionals had significantly higher odds of health insurance coverage (OR=1.78; p<0.001) compared to the unemployed. Respondents who reported manual labor employment were associated with even higher odds (OR=3.26; p<0.001) of coverage, while agricultural respondents were not significantly associated with coverage (OR=1.10; p<0.712) compared to the unemployed respondents. 2016 showed significantly higher odds of insurance coverage among the employed categories of professional (OR=1.93; p<0.001), agricultural (OR=3.19; p<0.001), and manual job holders (OR=2.62; p<0.001) when compared to the unemployed respondents.

Wealth was divided into five quantiles from poorest to richest and was associated with having insurance coverage in both years. The richest group in 2011 was associated with 17 times the likelihood of having health insurance (OR=17.10; p<0.001), and the rich group was associated with four times the odds of having health insurance (OR=4.34; p<0.001) compared to the poorest category of respondents. The poorer and middle groups were not statistically significantly different that the poorest group. In 2016, all other levels of wealth were associated with statistically significantly higher odds of having health insurance coverage compared to the poorest level. The poorer category had an OR of 2.43 (p<0.001), and the middle category had an OR of 4.00 (p<0.001). The rich group had an OR of 3.65 (p<0.001), and the richest group had an OR of 2.37 (p<0.001).

Newspaper exposure was statistically significantly associated with health insurance coverage in 2011 and 2016 compared to people with no exposure. This association also increased with increased frequency of exposure to newspapers. In 2011, the OR for newspaper exposure for less than once a week was 1.34 (p=0.003) in comparison to no exposure to newspaper. This increased for exposure of at least once a week to 1.80 (p<0.001). Similarly, in 2016, the OR for newspaper exposure for less than once a week was 1.27 (p=0.006). This increased for exposure at least once a week to 1.86 (p<0.001). Being exposed to television was associated with being covered by health insurance in both years. In 2011, the OR for less than once a week was 1.71 (p=0.013), and the OR for at least once a week was 1.83 (p=0.006). In 2016, the OR for less than once a week was 1.43 (p=0.013), and the OR for at least once a week was 1.25 (p=0.026) in comparison to no television exposure. In both years studied, when controlling for other variables, listening to the radio was not statistically significantly associated with health insurance coverage.

## Discussion

Findings of this study show that in 2011 and 2016 health insurance coverage tends to increase with higher education, older age, employment in professional or manual labor positions (when compared to the unemployed), greater wealth, newspaper exposure, and TV exposure. Even with these trends, while health insurance more than tripled between the two survey years; they remained low at 4.89% in 2016. In 2010/11, CBHI was piloted in 13 districts that were in four regions that mainly targeted rural areas. Expansion to other districts did not occur for a few years. Other studies have found that CBHI rates were much higher than SHI despite the SHI scheme being non-voluntary in principle (Atafu & Kwon, 2018; Ethiopian Health Insurance Agency, 2015). This indicates the SHI has not been effectively implemented to cover the formal sector. The low coverage rate of SHI and rural targets indicated that rural residents were more likely to be covered in 2011. Our results showed that in 2011, rural residents were more likely to be covered while in 2016, there was no significant difference among rural and urban residents. This suggests that CBHI schemes may have been active in some urban markets even though it was initially implemented in rural areas.

In both years studied, any form of exposure to television (TV) and newspaper were associated with having health insurance coverage compared to no exposure. However, there was no significant association with radio exposure. These results provide valuable information for policy makers as they move forward in their attempts to provide more people with health insurance coverage. Our analysis indicates that TV and newspapers were more effective outreach mechanisms, at least with early adopters, illustrating that their use could be more influential in the process of disseminating information about insurance programs. Ethiopia’s national communications campaign, which was launched to spread information about enrolling in the CBHI program, utilized journalists and television news reports to gather testimonies and propagate interest in the insurance program (Health Finance & Governance, 2014). In many developing countries, radio is the most effective form of mass media exposure (Kansanga et al., 2018). The lack of association between radio and health insurance coverage suggests that the current radio strategies have not yet been effectively executed, possibly due to the many different languages found across very large rural areas (Mohammed, 2013). One study that used different forms of communication in Ethiopia to disseminate information about child feeding practices found that a significant number of Ethiopians were not in areas reachable by radio broadcast, and therefore used mobile vans with speakers to spread their program (Kim et al., 2019).

The regression results also show that populations older than the reference group (15-24 years) were more likely to be covered by health insurance in both of the years studied. Yet the impact of older age decreased from 2011 to 2016. The age group reporting the highest rates of insurance was those 45 years and older, indicating that middle-aged people were more likely to have coverage. To advance insurance rates in Ethiopia over the long term, greater attention to younger age groups may be necessary. Younger individuals tend to be healthier and any individual contributions to the insurance system could aid in keeping insurance schemes solvent. Improving coverage among younger populations is also vital as health and health behaviors that begin earlier, potentially including insurance decisions, often continue into adult life. Ultimately, adolescent health is pivotal for the development of a nation due to its integral role in driving political and financial factors for the future of that nation’s endeavors (Viner et al., 2012).

Enrolling vulnerable populations in Ethiopia remains a challenge, though some progress has been made. Higher education doubled the chances of being insured even in 2016, though it had an even larger impact (OR=5.61, p<0.001) in 2011. Targeting vulnerable populations, including those with lower education levels, should be a priority for Ethiopia to ensure higher coverage rates. In 2016, all the wealth quintiles in comparison to the poorest group were statistically significant. The richest group was about 17 times more likely to have health insurance in 2011, which dropped to 2.37 times the likelihood in 2016. Over the years, wealth became less important as CBHI can be targeted towards poor and otherwise vulnerable populations. A similar trend was observed for occupation in which agricultural workers who were not likely to be enrolled in 2011 (OR=1.10, p<0.712), became more likely to have health insurance in 2016 (OR=3.19, p<0.001) compared to the unemployed population. This is a result of the rural focus of the CBHI scheme. Overall, enrollment in CBHI can be credited with making health insurance coverage more accessible. Although the overall enrollment rates remain low, Ethiopia is progressing with making insurance coverage more equitable.

In Ethiopia coverage remains limited, even while other countries in sub-Saharan Africa have achieved strong gains in the past few decades. Ghana has implemented a National Health Insurance Scheme (NHIS) since 2005, where enrollment increased from 6% to 38% from 2005 to 2014 (Aryeetey et al., 2016). Ghana’s NHIS has managed to adapt different models in order to fit Ghana’s needs. It uses a SHI model, though Ghana’s includes the informal sector unlike Ethiopia. It also uses CBHI-like entities with centralized authority and a National Health Insurance Fund to better provide coverage and financial sustainability (Odeyemi & Nixon, 2013). Similarly, the CBHI implemented in Rwanda has achieved over 80% coverage (Chemouni, 2018). Rwanda is the only country in sub-Saharan Africa to have attained such high coverage. Their health system overall is financed by state funds as well as individual donors. Rwanda’s government plays a role in paying for the premium and copayments for indigent and vulnerable populations (Odeyemi, 2014). Both countries are successful due to financing by the government, the elimination or reduction of copayments, and uniform and comprehensive benefit packages (Chemouni, 2018; Odeyemi, 2014; Odeyemi & Nixon, 2013). Applying some of these lessons to Ethiopia will require significant political will and financial investment.

## Limitations

One limitation of this study is that cross sectional data is used so causality claims cannot be made. Second, the dataset does not categorize what type of insurance respondents have whether it be SHI or CBHI. Although SHI is supposed to be mandatory, many eligible individuals have not enrolled. SHI rates remain low, and most of the growth in coverage likely reflects growth in CBHI. Third, it is not known to what extent the government emphasized health insurance through different media mechanisms in different regions, and respondents were not asked how they had heard about insurance programs. Thus, this study uses access to media as a proxy measure.

## Conclusion

Health insurance coverage in Ethiopia increased 3.30 times from 2011-2016, but their overall coverage level remains significantly low at 4.89%. More than 90% of males and females remain uninsured. Pairing these results with data collected on the success of Ethiopia’s Community-Based Health Insurance could help identify approaches for targeting the large populations that remain uninsured. The comparison of results over the five-year period reveals where insurance rates have improved as well as the populations that are at risk for being left behind. As Ethiopia continues to grow economically, more widespread access to healthcare and health insurance could prove crucial in ensuring a healthy workforce, providing for additional future economic gains while securing the right to health.

## Data Availability

Data is available upon request from the Demographic Health Survey program website.

https://dhsprogram.com/Countries/Country-Main.cfm?ctry_id=65&c=Ethiopia&Country=Ethiopia&cn=&r=1

